# A Novel Protein Drug, Novaferon, as the Potential Antiviral Drug for COVID-19

**DOI:** 10.1101/2020.04.24.20077735

**Authors:** Fang Zheng, Yanwen Zhou, Zhiguo Zhou, Fei Ye, Baoying Huang, Yaxiong Huang, Jing Ma, Qi Zuo, Xin Tan, Jun Xie, Peihua Niu, Wenlong Wang, Yun Xu, Feng Peng, Ning Zhou, Chunlin Cai, Wei Tang, Xinqiang Xiao, Yi Li, Zhiguang Zhou, Yongfang Jiang, Yuanlin Xie, Wenjie Tan, Guozhong Gong

**Author notes:** Correspondence to: Guozhong Gong, Department of Infectious Diseases, the Second Xiangya Hospital, Central South University, Changsha, Hunan 410011, China. or Wenjie Tan, National Institute for Viral Disease Control and Prevention, Chinese Center for Disease Control and Prevention, Beijing 102206, China. or Yuanlin Xie, Department of Infectious Diseases, the First Hospital of Changsha, Changsha, Hunan 410011, China. or Yongfang Jiang, Department of Infectious Diseases, the Second Xiangya Hospital, Central South University, Changsha, Hunan 410011. equal work (Fang Zheng, Yanwen Zhou, Zhiguo Zhou, Fei Ye).

## Abstract

**Background:** Novaferon, a novel protein drug approved for the treatment of chronic hepatitis B in China, exhibits potent antiviral activities. We aimed to determine the anti-SARS-CoV-2 effects of Novaferon in vitro, and conducted a randomized, open-label, parallel group study to explore the antiviral effects of Novaferon for COVID-19.

**Methods:** In laboratory, the inhibition of Novaferon on viral replication in cells infected with SARS-CoV-2, and on SARS-CoV-2 entry into healthy cells was determined. Antiviral effects of Novaferon were evaluated in COVID-19 patients with treatment of Novaferon, Novaferon plus Lopinavir/Ritonavir, or Lopinavir/Ritonavir. The primary endpoint was the SARS-CoV-2 clearance rates on day 6 of treatment, and the secondary endpoint was the time to the SARS-CoV-2 clearance in COVID-19 patients

**Results:** Novaferon inhibited the viral replication in infected cells (EC_50_=1.02 ng/ml), and protected healthy cells from SARS-CoV-2 infection (EC_50_=0.1 ng/ml). Results from the 89 enrolled COVID-19 patients showed that both Novaferon and Novaferon plus Lopinavir/Ritonavir groups had significantly higher SARS-CoV-2 clearance rates on day 6 than the Lopinavir/Ritonavir group (50.0% vs.24.1%, p = 0.0400, and 60.0% vs.24.1%, p = 0.0053). Median time to SARS-CoV-2 clearance were 6 days, 6 days, and 9 days for three groups respectively, suggesting a 3-dayreduction of time to SARS-CoV-2 clearance in both Novaferon and Novaferon plus Lopinavir/Ritonavir groups compared with Lopinavir/Ritonavir group.

**Conclusions:** Novaferon exhibited anti-SARS-CoV-2 effects in vitro and in COVID-19 patients. These data justified the further evaluation of Novaferon.

## 1. Introduction

COVID-19 by the infection of a novel coronavirus, SARS-CoV-2, has become a global pandemic and caused more than1,000,000 confirmed cases and over 60,000 deaths [1-4]. Effective antiviral drugs for COVID-19 might lead to earlier clearance of SARS-CoV-2 that in turn helps to slow down or stop the progress of disease course for patients. The elimination of virus in patients at early stage would also contribute to the reduction of transmission of this deadly virus. With immediate availability and established safety profiles, approved antiviral drugs were targeted with the desire to find effective anti-SARS-CoV-2 drugs in the shortest time possible [5]. However, none of the most promising antiviral drugs for COVID-19 has been proved effective yet. Most published findings for the antiviral treatment of COVID-19 were based on the individual case reports or cellular antiviral results [6,7]. Despite the lack of convincing evidence, Lopinavir/Ritonavir was quickly selected and recommended as an antiviral drug for COVID-19 in China since January. So far, only limited observations of Lopinavir/Ritonavir for coronavirus in SARS patients were reported[8]. Results from a recently completed trial of Lopinavir/Ritonavir in patients with severe COVID-19 generated disappointed outcomes and showed no significant antiviral effects[9]. Without effective antiviral drugs for COVID-19, health care workers have no alternative choices but mainly rely on the supportive and symptomatic treatments to manage COVID-19 patients. The rapid spread and daily increase of large death numbers in over 100countries perhaps represent one of the biggest medical and humanity challenges, it is even more critical than two months ago to find antiviral drugs with clinical evidence for COVID-19 patients[10,11].

Novaferon, a novel antiviral protein drug which has been approved for the treatment of chronic hepatitis B in China and exhibited broad-spectrum antiviral properties (unpublished data, available on requests), became an obvious candidate as a potential antiviral drug for COVID-19. Novaferon consists of 167 amino acids and is not a naturally existing protein. According to the published information in a US patent (US 7,625,555 B2), this novel protein was invented in laboratory on the technical basis of DNA shuffling technology and named as Novaferon by its inventors[12]. Novaferon has similar properties of human interferon, but its antiviral activities were greatly improved and are at least 10 times more potent than human interferon alpha-2b[13]. The antiviral efficacy of Novaferon was demonstrated by clinical studies conducted in China. In April 2018, Novaferon was approved in China by former CFDA (Chinese Food and Drug Administration) for the treatment of chronic hepatitis B. Novaferon protein’s non-proprietary name was temporarily defined as “recombinant cytokine gene-derived protein injection” by Chinese Pharmacopeia Committee, and the recommended international non-proprietary name (rINN) by WHO is not available yet. For convenience purposes, Novaferon was used as the drug name in our study.

In the present study, we specifically focused on observing the antiviral effects of Novaferon. We first determined whether Novaferon was able to inhibit SARS-CoV-2 at cellular level, and then conducted a randomized, open-label, parallel group trial to explore the antiviral effects of Novaferon in patients with COVID-19 by observing the SARS-CoV-2 clearance rates at different times during the treatment period. The primary endpoint was the SARS-CoV-2 clearance rates on day 6, and the secondary endpoint was the median time to reach SARS-CoV-2 clearance in patients with COVID-19. As a popular and standard antiviral drug for COVID-19 in China, Lopinavir/Ritonavir was included in this study to serve as a control for comparison.

## 2. Methods

### In vitro antiviral testes

All the in vitro experiments were conducted in a biosafety level-3 (BSL3/P3) laboratory at Chinese Center for Disease Control and Prevention (Chinese CDC). We first determined whether Novaferon inhibited the SARS-CoV-2 replication within the Vero E6 cells which were already infected with SARS-CoV-2. Vero E6 cells (African green monkey kidney cells) were obtained from American Type Culture Collection (ATCC, Manassas, VA, USA) and maintained in Modified Eagle Medium (MEM; Gibco Invitrogen, Carlsbad, CA, USA) with 10% fetal bovine serum (FBS; Thermo Scientific HyClone, South Logan, UT) at37 □in the incubator with 5% CO_2_. A clinically-isolated strain of SARS-CoV-2 (C-Tan-nCov Wuhan strain 01) was propagated in Vero E6 cells before the experiments, and the plaque assay was used to quantify the titre (plaque forming unit, PFU) of the SARS-CoV-2. Blank Vero E6 cells in 96-well plates with a density of 1 × 10^4^per well were incubated with C-Tan-nCov Wuhan strain 01 (100PFU) for 2 hours to induce the infection of the cultured cells by SARS-CoV-2. After virus-containing supernatants were removed and cells rinsed, medium with various concentrations of Novaferon(0, 0.001, 0.01, 0.1, 1,10, or 100ng/ml)was added and the cells were then incubated for 24 hours to allow the viral replication. After 24 hours, 100 μL of supernatants from each well were collected, and the total viral RNA was extracted (Full-automatic nucleic acid Extraction System from TIANLONG) and measured. Takara Bio’s One Step Prime Script Real-time PCR (RT-PCR) Kit (Perfect Real Time) was used to detect the copies of the virus RNA.

The Quantitative real-time PCR cycle threshold (Ct) was obtained, and the inhibition rates of virus replication by each Novaferon concentration were calculated. The Ct number of controls obtained in the absence of Novarferon was 22.6 and considered as 100%. The Ct numbers from SARS-CoV-2 infected Vero E6 cells with the addition of various concentrations of Novaferon were measured to calculate the inhibition percentages. The half-maximal effective concentration (EC_50_) was calculated. The cytotoxicity of Novaferon on Vero E6 cells was assessed, and the half-maximal cytotoxic concentration (CC_50_) of Novaferon on Vero Cells was determined by observing the cytopathic effects (CPE) of Novaferon. The selectivity index (CC_50_/EC_50_) was then calculated.

We further observed whether the previous treatment of Vero E6 cells with Novaferon protected the cells from viral entry through exposure of the pre-treated cells to SARS-CoV-2 later. Detailed operation procedures were identical to the above description, except that the step orders were changed to allow the observation of the preventive effects of Novaferon. Briefly, blank Vero E6 cells were incubated with series concentrations of Novaferon for 2 hours, and the supernatants containing Novaferon were then removed. The pre-treated Vero E6 cells were exposed to SARS-CoV-2 by incubation with C-Tan-nCov Wuhan strain 01 (100PFU) for 2 hours, and the supernatants containing SARS-CoV-2 were then removed. Fresh medium was added, and the cells were incubated for 48 hours. 100 μL of supernatants were taken from each well, and the total viral RNA in the supernatants was measured using the same methods described above. The Ct number obtained from Vero E6 cells without pre-treatment of Novaferon was considered as 100%, and the decreased Ct numbers obtained from the pre-treated Vero E6 cells with various concentrations of Novaferon were used to calculate the inhibition percentages. The preventive effects of Novaferon were then determined by observing the viral RNA reduction inthe cells pre-treated with Novaferon. The EC_50_ of Novaferon for the observed preventive effects was decided accordingly.

### Laboratory detection of SARS-CoV-2 nucleic acids by RT-PCR in nasopharyngeal swab samples

SARS-CoV-2 virus nucleic acids were detected by RT-PCR using SLAN-96P automatic medical PCR analysis system. The SARS-CoV-2 nucleic acid detection kit was obtained from Sensure Biotechnology Co. Ltd(Hunan province, China), which has been approved for clinical test of SARS-CoV-2 by NMPA (National Medical Products Administration). Procedures of the tests strictly followed the protocol of the kit. Samples from nasopharyngeal swab were collected in accordance with the standard procedures of the New Coronavirus Infection Pneumonia Laboratory Test Guide. FAM (ORF-1ab region) was used as fluorescent detection channel, and ROX (N gene) channels were used to detect the SARS-CoV-2 nucleic acids, while HEX channel was used as the internal standard. Cycle parameter steps were in the following order: 1)reverse transcription at 50□ for 30 minutes for 1 cycle; 2) pre-denaturation of cDNA at 95□ for 1 minute for 1 cycle; 3) denaturation at 95□ for 15 seconds, and annealing, extension and fluorescence acquisition at 60□ for 30 seconds for 45 cycles. 4) cooling at 25□ for 10 seconds for 1 cycle. Positive results were determined by comparing between the Ct numbers of the testing samples and the standard Ct number that was 40 in this assay.

### Clinical Study

#### Patients

The study was originally designed as a multi-center study across hospitals in Changsha city and in other cities of Hunan Province, China. However, per government order, all patients from hospitals in Changsha city had to be relocated to the First Hospital of Changsha, a designated treatment center for all COVID-19 patients in Changsha city, and hospitals in other cities of Hunan Province were not able to participate due to various reasons. The study was changed to a single center study. This study was approved by the ethics committee of the First Hospital of Changsha (file number KX-2020002) andwas conducted at the hospital. The study was also registered at the Chinese Clinical Trial Registry (http://www.chictr.org.cn/), number ChiCTR2000029496.

Hospitalized COVID-19 patients with confirmed SARS-CoV-2 detection, clinically classified as moderate or severe, at the age over 18 years, and without comorbidity of severe heart, lung, brain diseases, were eligible for enrolling into this study. Moderate patients were defined as “patients with fever, symptoms of respiratory system and pneumonia changes in CT images, and severe patients were defined as “patients with any of the following: ⍰ Respiratory distress, respiratory frequency ≥30/minute;⍰ Under rest status, arterial oxygen saturation (SaO_2_)≤ 93%;⍰Arterial partial pressure of oxygen(PaO_2_)/fraction of inspired oxygen (FiO_2_)≤ 300mmHg. In this study, we aimed to observe the moderate and severe COVID-19 patients as these patients would likely benefit more from antiviral treatments.

### Trial design and treatments

This was a randomized, open-label, parallel group study. Patients eligible for the study were assigned, in a 1:1:1 ratio, to Novaferon, Novaferon plus Lopinavir/Ritonavir, or Lopinavir/Ritonavir group. A SAS generated simple randomization schedule was prepared by a statistician not involved in the trial. Based on the sequence that patients enrolled into the study, the patients were assigned to a treatment group which was implemented by a research assistant. Informed consents were obtained from all enrolled patients.

Antiviral effects were assessed on day 3, day 6, and day 9 after starting drug administration. For the above antiviral drugs involved in the study, Novaferon (temporary non-proprietary name: recombinant cytokine gene-derived protein injection) was manufactured in Qingdao city of Shandong Province by Genova Biotech (Qingdao) Company Limited. The approved dosage of Novaferon for hepatitis B application is the daily injection of 10 μg of protein in 1.0ml volume per vial. Lopinavir/Ritonavir (Kaletra) was manufactured by AbbVie Inc. and each tablet contained 200mg of Lopinavir and 50mg of Ritonavir.

The total daily doses (40 μg) of Novaferon were administered to patients twice per day by the oxygen-driven atomized inhalation for 15 minutes of 20 μg of Novaferon (2 × l ml vials) diluted with saline. For patients receiving Lopinavir/Ritonavir (Kaletra), 2 tablets were orally taken twice per day.

### Assessments

Samples of nasopharyngeal swab on day 3, day 6 and day 9 during the 7 to 10-day course of antiviral treatment were collected from the patients and tested for SARS-CoV-2 nucleic acids by RT-PCR. The negative conversion of SARS-CoV-2 in nasopharyngeal swab samples was defined as the SARS-CoV-2 clearance in COVID-19 patients. Adverse events were monitored throughout the trial, reported and graded based on WHO Toxicity Grading Scale for Determining the Severity.

The peak levels of SARS virus were around 10 days after onset and then the viral level began to decrease without effective antiviral treatment in SARS patients[14]. Considering the homology of gene sequences of SARS-CoV-2 and SARS was over 90%[2], we assumed that the intervention of antiviral drugs in COVID-19 patients would likely enhance or shorten the time of viral clearance. In this regarding, the primary endpoint for this study was decided as the SARS-CoV-2 clearance rates in COVID-19 patients assessed on day 6 of antiviral treatment. The secondary endpoint was the time till the SARS-CoV-2 clearance.

### Statistical analysis

Statistical analysis was performed on an intent-to-treat basis, and all patients randomized and treated at least once with the study medications were included for the primary analysis. For patient demographics information and baseline disease characteristics, qualitative variables were compared among treatment groups with the use of Chi-square test, and quantitative variables were compared with the use of an ANOVA model. Only the overall differences among the three treatment groups were tested (based on null hypothesis, “all three groups were the same”, against an alternative hypothesis, “at least one group was different”) and therefore, no pairwise comparison was performed for baseline characteristics.

For the primary endpoint, SARS-CoV-2 clearance rate, estimates of the rates were calculated based on a binomial distribution. Difference between treatment groups was tested using the Chi-square test. To control the overall significance level for the study, the three pairwise comparisons for the primary endpoint were performed at the two-sided alpha = 0.05 using a closed testing procedure according to the following order:□Novaferon plus Lopinavir/Ritonavir vs. Lopinavir/Ritonavir alone; □Novaferon alone vs. Lopinavir/Ritonavir alone; □Novaferon plus Lopinavir/Ritonavir vs. Novaferon alone. For the secondary endpoint, time to SARS-CoV-2 clearance, median time for each group was estimated with the use of the Kaplan–Meier method and treatment differences were tested using log-rank test. All tests were two-sided, with a p value of less than 0.05 considered to indicate statistical significance. Analysis was conducted using SAS V9.2. For missing SARS-CoV-2 clearance status, Last Observation Carried Forward (LOCF) analysis was presented as the primary analysis. For purpose of sensitivity analyses, complete case analysis and worst case imputation methods were also performed. For the worst case imputation, missing SARS-CoV-2 status was replaced with ‘positive’.

The planned sample size of 90 patients (30 patients per group) was not determined based on statistical consideration.

Adverse events were reported and graded using WHO Toxicity Grading Scale for determining the severity. Incidence of adverse events was summarized descriptively without a formal statistical test.

## 3. Results

### Inhibitory effects of Novaferon on SARS-CoV-2 at cellular level

As shown in fig.1, incubation of Novaferon (0.1□100ng/ml) with SARS-CoV-2-infected Vero E6 cells resulted in the dose-dependent reductions of the SARS-CoV-2 RNA that was released from the infected Vero E6 cells. The half-maximal effective concentration (EC_50_) of Novaferon was 1.02 ng/ml. The tested Novaferon concentrations showed minimal cytotoxicity to Vero E6cells, and the half-maximal cytotoxic concentration (CC_50_) was over 100ng/ml. The selectivity index (CC_50_/EC_50_) was over 98. These data indicated that Novaferon effectively inhibited the viral replication within SARS-CoV-2-infected cells. In addition, healthy Vero E6 cells that were previously treated with Novaferon obtained the ability of resisting the viral entry, as indicated by the reduction of viral RNA after exposure of the treated cells to SARS-CoV-2 later. Novaferon exhibited this preventive effect efficiently with the EC_50_(0.1 ng/ml), lower than the EC_50_ for inhibiting SARS-CoV-2 replication in the infected cells.

**Figure 1.**
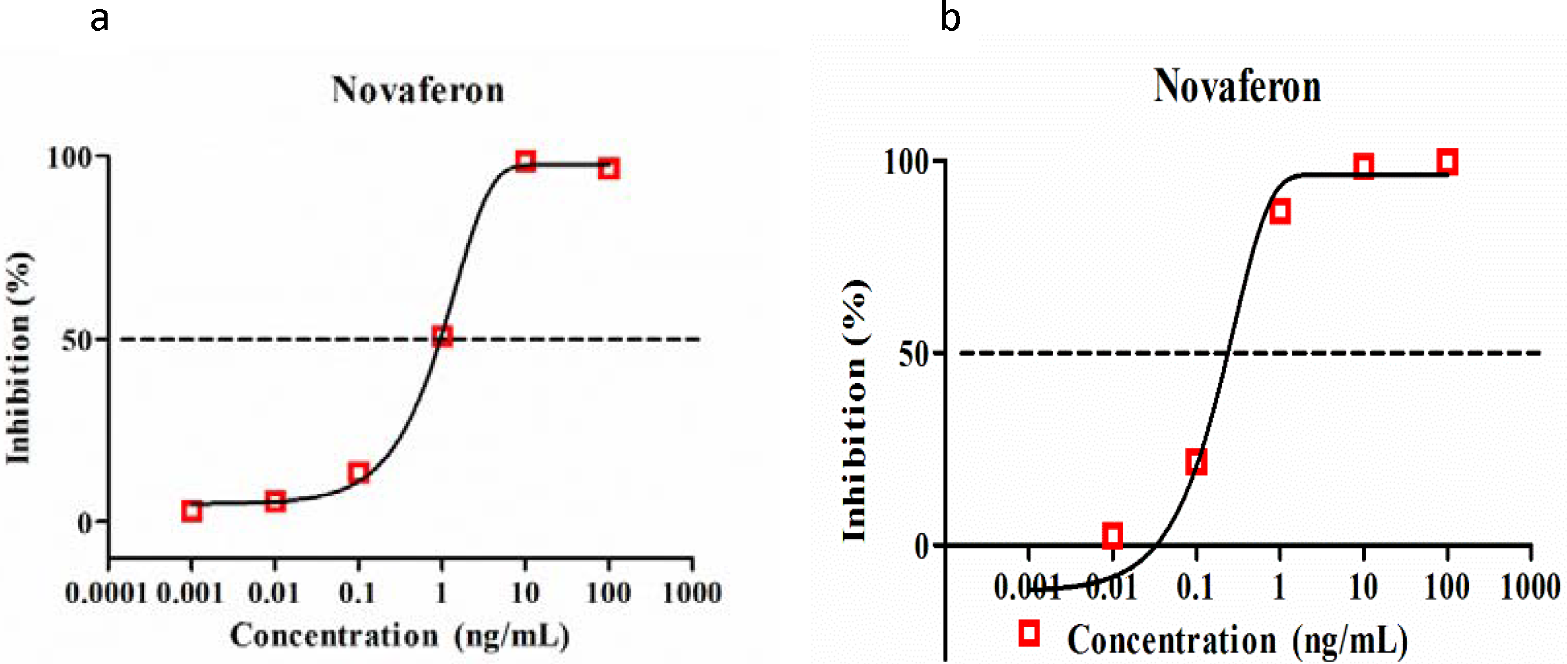
Concentration-effect relationship of the inhibition of Novaferon on SARS-CoV-2 replication. The experiments were done in triplicates. a. inhibitory effects of Novaferon after virus infection; b. inhibitory effects of Novaferon before virus infection.

These data suggested that Novaferon inhibited viral replication in SARS-CoV-2-infected cells and enabled healthy cells to resist the viral entry.

### Clinical study

#### Patients and Treatments

A total of 92 patients with moderate or severe illness were assessed for the eligibility criteria and 3 patients were excluded. 89 patients were randomized into the study from 1 February to 20 February 2020. Of the 89 patients, 30, 30, and 29 patients were assigned into Novaferon group, Novaferon plus Lopinavir/Ritonavir group or Lopinavir/Ritonavir group respectively. The baseline demographic and clinical characteristics of the 89 patients were shown in Table 1. Among the 89 patients, 84 were moderate ill and 5 severe ill. There were no apparent differences among the groups with respect to any of the patient demographics and the baseline characteristics.

**Table 1.** Patient Demographics and Baseline Disease Characteristics

| Statistics | Novaferon<br>N=30 | LPV/r*+Novaferon<br>N=30 | LPV/r<br>N=29 | p<br>value† |
| --- | --- | --- | --- | --- |
| Age |  |  |  | 0.0584 |
| Mean | 50.1 | 48.8 | 41.1 |  |
| Min | 29 | 21 | 18 |  |
| Median | 46.5 | 50.0 | 37.0 |  |
| Max | 76 | 74 | 73 |  |
| Gender |  |  |  | 0.4376 |
| Male | 17(56.7%) | 13(43.3%) | 12 (41.4%) |  |
| Female | 13(43.3%) | 17 (56.7%) | 17 (58.6%) |  |
| Disease stage |  |  |  | 0.8262 |
| Severe | 2 (6.7%%) | 2 (6.7%) | 1 (3.4%) |  |
| moderate | 28 (93.3%) | 28 (93.3%) | 28 (96.6%) |  |

\*LPV/ r: Lopinavir/Ritonavir.
†p value: Based on null hypothesis, “all three groups were the same”, against an alternative hypothesis, “at least one group was difference”; ‘sex’ and ‘disease stage’ were tested with the use of a Chi-square test, and ‘age’ with an ANOVA model;

#### Primary endpoint

Table 2 summarized the complete RT-PCR test results of all 89 patients on day 3, day 6, and day 9 after starting drug administration. The negative results of SARS-CoV-2 nucleic acid detection in the tested samples served as the indicator of in vivo SARS-CoV-2 clearance in patients. The SARS-CoV-2 clearance rates on day 3, day 6, and day 9 in three treatment groups were presented and compared (Table 2).On day 3, SARS-CoV-2 clearance rates were 16.7%(5/30) in Novaferon group, 36.7% (11/30) in Novaferon plus Lopinavir/Ritonavir group, and 10.3% (3/29) in Lopinavir/Ritonavir group respectively. SARS-CoV-2 clearance rate in Novaferon plus Lopinavir/Ritonavir group was significantly higher than in Lopinavir/Ritonavir group on day 3 (36.7%vs. 10.3%, p = 0.0175). No significant difference between Novaferon group and Novaferon plus Lopinavir/Ritonavir group was observed. On day 6, SARS-CoV-2 clearance rates in Novaferon group and Novaferon plus Lopinavir/Ritonavir group reached to 50.0% (15/30) and 60.0% (18/30) respectively, and were significantly higher than in Lopinavir/Ritonavir group (50.0% vs.24.1%, p = 0.0400, and 60.0% vs.24.1%, p = 0.0053, respectively). There was no statistically significant difference between Novaferon group and Novaferon plus Lopinavir/Ritonavir group, suggesting the similar extents of enhancedSARS-CoV-2 clearance on day 6 by Novaferon alone or together with Lopinavir/Ritonavir. On day 9, SARS-CoV-2 clearance rates were 56.7% (17/30) in Novaferon group, 70.0% (21/30) in Novaferon plus Lopinavir/Ritonavir group, and 51.7% (15/29) in Lopinavir/Ritonavir group. There were no statistically significant differences between the groups.

**Table 2.** Summary of SARS-CoV-2 clearance rates

|  |  |  |  | p value† |  |  |
| --- | --- | --- | --- | --- | --- | --- |
|  | Novaferon<br>(N=30) | LPV/r*+<br>Novaferon<br>(N=30) | LPV/r<br>(N=29) | LPV/r<br>vs.<br>Novaferon | LPV/r<br>vs. LPV/r +<br>Novaferon | Novaferon<br>vs. LPV/r +<br>Novaferon |
| Day 3 | 16.7%<br>(5/30) | 36.7%<br>(11/30) | 10.3%<br>(3/29) | 0.4783 | 0.0175 | 0.0798 |
| Day 6 | 50.0%<br>(15/30) | 60.0%<br>(18/30) | 24.1%<br>(7/29) | 0.0400 | 0.0053 | 0.4363 |
| Day 9 | 56.7%<br>(17/30) | 70.0%<br>(21/30) | 51.7%<br>(15/29) | 0.7032 | 0.1502 | 0.2839 |

\*LPV/ r: Lopinavir/Ritonavir.
†p values for comparisons between treatment groups using Chi-square test;At any visit, missing viral RNA outcome was imputedwith Last Observation Carried Forward (LOCF) method.

#### Secondary endpoint

The median time to SARS-CoV-2 clearance were 6 days, 6 days, and 9 days for Novaferon group, Novaferon plus Lopinavir/Ritonavir group, and Lopinavir/Ritonavir group respectively, indicating a 3-day reduction of time for reachingSARS-CoV-2 clearance in both Novaferon and Novaferon plus Lopinavir/Ritonavir groups comparing with Lopinavir/Ritonavir group (Table 3).During the observation period, none of the moderate ill patients in Novaferon group and Novaferon plus Lopinavir/Ritonavir group progressed to severe ill. In contrast, 4 moderate ill patients in Lopinavir/Ritonavir group progressed to severe ill.

**Table 3.**
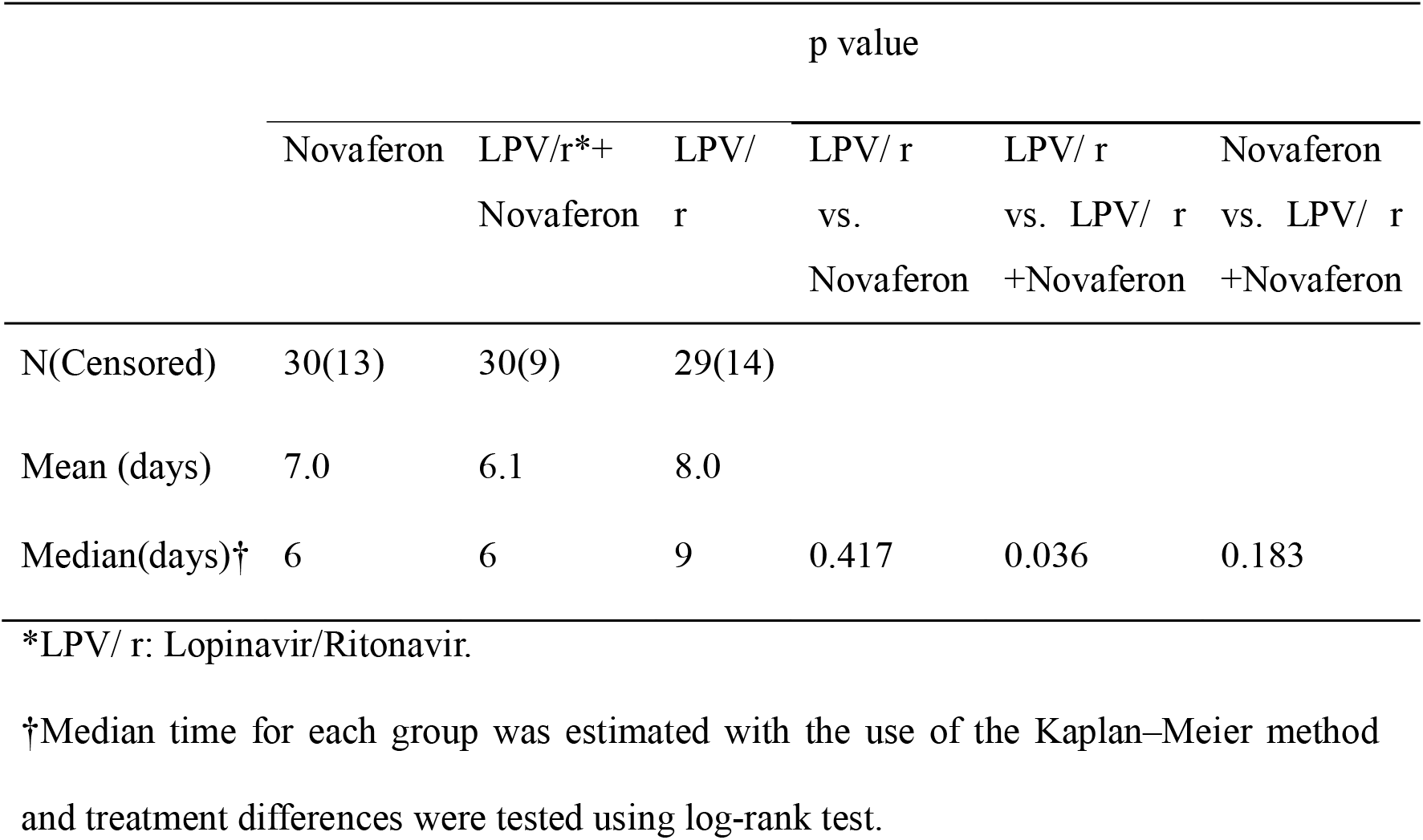
Analysis for Time to SARS-CoV-2 Clearance

#### Sensitivity analysis for missing data

Analyses based on both complete case analysis and worst case imputationforSARS-CoV-2 clearance rates showed little differences with the LOCF analysis, and the statistical conclusions for all the treatment comparisons remained the same.

#### Adverse effects

No severe adverse events (SAE) associated with the tested antiviral drugs were observed. Reported grade1, and grade 2 adverse events (AE) were described below. 2 patients in Lopinavir/Ritonavir group had diarrhea. 1 patient in Novaferon plus Lopinavir/Ritonavir group had symptoms of nausea, vomiting and diarrhea and 1 patient in Novaferon plus Lopinavir/Ritonavir group had transient, slight elevation of blood urea nitrogen (BUN). Transient, slight elevation of alanine aminotransferase (ALT) occurred in 2 patients in Lopinavir/Ritonavir group and 1 patient in Novaferon plus Lopinavir/Ritonavir group. Without extra interventions and termination of antiviral treatment, all the transient elevation of BUN and ALT returned to the normal levels during the observation period.

## 4. Discussion

No matter whether exhibiting good, poor or none anti-COVID-19 effects, Lopinavir/Ritonavir in this study served as the control and allowed us to assess the antiviral effects of Novaferon by analyzing the differences between Novaferon group and Lopinavir/Ritonavir group. In this regarding, the significantly higher SARS-CoV-2 clearance rates on day 6, as the primary endpoint, observed in patients with the treatment of Novaferon alone or together with Lopinavir/Ritonavir comparing with Lopinavir/Ritonavir alone, indicated that Novaferon indeed exhibited antiviral effects for COVID-19 patients. The 3-day reduction of time to SARS-CoV-2 clearance in patients with Novaferon treatment further supported the antiviral effects of Novaferon.

As viral shedding in the early stages of COVID-19 represents a major challenge for controlling the transmission of SARS-CoV-2[15], an effective antiviral drug capable of eliminating the virus in the quickest time would not only contribute to earlier recovery of COVID-19 patients but also to the reduction of virus transmission by early stage patients who were reported with the highest viral loads[15].

The antiviral effects of Novaferon for COVID-19 patients were in consistence with the laboratory findings. Inhibition of the viral replication by Novaferon at cellular level was very efficient as indicated by the low EC_50_(1.02 ng/ml). More interestingly, healthy cells that were pre-treated by Novaferon obtained the ability, in the absence of Novaferon, to resist the viral entry into cells when the treated cells were exposed to SARS-CoV-2 later (EC_50_ 0.1 ng/ml). It might be worth to explore the potential use of Novaferon as a preventive agent for high risk population, especially for health care workers who have to routinely contact COVID-19 patients.

The viral loads in COVID-19 patients were reported to reach peak levels around 5□6 days after onset [16], and for severe patients,the average time from onset to severe stage took about one week[17]. The early clearance of SARS-CoV-2 might help to shorten the disease course or to prevent the disease progress in patients with moderate COVID-19. Considering the high viral loads and peak levels of SARS-CoV-2 were found in first week of onset[15,16], the increased SARS-CoV-2 clearance rates on day 6 in COVID-19 patients with Novaferon treatment were unlikely related to the natural disease course. Rather, the observed enhancement of SARS-CoV-2 clearance apparently reflected the antiviral effects of Novaferon in observed COVID-19 patients.

### Limitations of this study

Our study has several limitations. First, all observations were done at one hospital in one city. Second, the sample size was relatively small and was not based on statistical consideration as limited by the availability of COVID-19 patients in Changsha city. Third, the unexpected difficulties associated with the COVID-19 outbreak compromised the quality of this study. For example, it’s highly possible that adverse events were under-reported due to the lack of medical staff and the risk situation. However, these limitations shouldn’t change the overall conclusion for thisrandomized trial because the antiviral assessments were strictly performed according to a vigorous standard.

## 5. Conclusions

Novaferon exhibited anti-SARS-CoV-2 effects at cellular level and in patients with COVID-19. Data obtained from this randomized, open-label, parallel group trial preliminarily demonstrated the anti-SARS-CoV-2 effects of Novaferon for COVID-19, and justified large-scale clinical studies to verify the efficacy of Novaferon. Novaferon might be considered as a potential antiviral drug for COVID-19.

## Data Availability

After formal publication, data related to study protocol, statistical analysis plan, etc. will be made available to others with reasonable written request to corresponding authors. The requests will be evaluated, and data provided on the basis of reasonability, detailed study design and protocol, permission of legal and government rules, and approval from the study participating institutes.

## Acknowledgments

We thank all the medical and management staff, who came from hospitals across Changsha City and worked at the First Hospital of Changsha, for their courage and dedication to COVID-19 patient care and overall operations of the hospital during the difficulty time. We especially thank Dr. Charlie Chen of SRD ClinMax Corporation for conducting the statistical analysis.

## Author Contributors

Drs F Zheng, Y Zhou, Z Zhou and F Ye contributed equally to this article and share first authorship. Drs Y Jiang, YXie, W Tan, GGong should be considered as co-correspondence authors, have full access to all the data in the study and take responsibility for the integrity and accuracy of the data.

## Ethical approval

The study protocol was approved by the ethics committee of First Hospital of Changsha.

